# EFFICACY OF INTRAVENOUS IMMUNOGLOBULIN ALONE ON CORONARY ARTERY LESION REDUCTION IN KAWASAKI DISEASE

**DOI:** 10.1101/2024.07.11.24310310

**Authors:** Ho-Chang Kuo, Ming-Chih Lin, Chung-Chih Kao, Ken-Pen Weng, Yun Ding, Chih-Jung Chen, Sheng-Ling Jan, Kuang-Jen Chien, Chun-Hsiang Ko, Chien-Yu Lin, Wei-Te Lei, Ling-Sai Chang, Mindy Ming-Huey Guo, Kuender D. Yang, Karl G. Sylvester, Zhi Han, John C. Whitin, Lu Tian, Henry Chubb, Scott R. Ceresnak, Doff McElhinney, Harvey J. Cohen, Xuefeng B. Ling

## Abstract

**Background:** Though Aspirin and intravenous immunoglobulin (IVIG) remain the standard treatments for Kawasaki Disease (KD) to minimize coronary artery damage, the duration and dosage of aspirin are inconsistent across hospitals. However, the lack of multi-center randomized trials prevents definitive answers to the impact of high-dose aspirin.

**Methods:** This clinical trial was structured as a prospective, evaluator-blinded, multi-center randomized controlled trial with two parallel arms, aiming to assess the effectiveness of IVIG as a standalone primary therapy of KD in comparison to the combination of IVIG with high-dose aspirin therapy. KD patients were enrolled between September, 2016 and August, 2019. A final cohort of 134 patients were randomly assigned to the standard and test groups with 69 and 65 patients, respectively. The Standard group received IVIG (2 g/kg) along with aspirin (80-100 mg/kg/day) until fever subsided for 48 hours. The test group received IVIG (2 g/kg) alone. Following the initial treatment, both groups received a daily aspirin dose (3-5 mg/kg) for six weeks. The primary outcome measure was the occurrence of coronary artery lesions (CAL) at the 6-8 weeks mark. The secondary outcome is IVIG resistance.

**Results:** The overall rate of CAL in test group decreased from 10.8% at diagnosis to 1.5% and 3.1% at 6 weeks and 6 months, respectively. The CAL rate of standard group declined from 13.0% to 2.9% and 1.4%, with no statistically significant difference (P>0.1) in the frequency of CAL between the two groups. Furthermore, no statistically significant differences were found for treatment (P>0.1) and prevention (P>0.1) effect between the two groups.

**Conclusions:** This marks the first prospective multi-center randomized controlled trial comparing the standard treatment of KD using IVIG plus high-dose aspirin against IVIG alone. Our analysis indicates that addition of high-dose aspirin during initial IVIG treatment is neither statistically significant nor clinically meaningful for CAL reduction.

**Registration:** URL: http://www.clinicaltrials.gov; identifier: NCT02951234

**What is New?:**

- This study represents the first multi-center randomized controlled trial investigating the efficacy of high-dose aspirin or intravenous immunoglobulin (IVIG) during the acute stage of KD. This study assessed the impact of discontinuing high-dose aspirin (80–100 mg/kg/day) on the occurrence of CAL during the acute phase treatment of Kawasaki Disease.
- No significant differences were observed between high-dose aspirin plus IVIG treatment and IVIG alone treatment in terms of the frequency of abnormal coronary artery abnormalities. Additionally, our analysis revealed no statistically significant differences in either the treatment effect (the number of cases successfully treated) or prevention effect (the prevention of new cases) between these two treatments.

**What Are the Clinical Implications?:**

- Comparison analysis indicated the non-inferiority between two groups with or without high-dose aspirin.
- Administering the standard 2 g/kg/day IVIG without high-dose aspirin (80–100 mg/kg/day) during the acute phase therapy for KD does not increase the risk of coronary artery lesions, which are a primary cause of morbidity and mortality in KD patients.
- Addition of high-dose aspirin during initial IVIG treatment is not statistically significant or clinically meaningful.

## Introduction

Kawasaki disease (KD) is an acute vasculitis characterized by self-limiting tendencies, predominantly impacting medium-sized vessels and resulting in coronary artery lesions^1, 2^ such as dilatations, fistulas, infarctions^3^ and aneurysms^4^. Initial treatment typically includes intravenous immunoglobulin (IVIG) and high-dose aspirin (80–100 mg/kg/day), which synergistically exert anti-inflammatory effects according to the American Heart Association (AHA) criteria. It’s noteworthy that high-dose aspirin, with its anti-inflammatory properties, primarily addresses inflammation, while lower doses are primarily directed at thrombosis.

The commonly accepted understanding of CAL, also known as coronary artery abnormalities (CAA), is based on diagnostic criteria set by the Japanese Ministry of Health. These criteria involve a maximum internal diameter exceeding 3 mm in children under 5 years old or exceeding 4 mm in children aged 5 years and above. Furthermore, the criteria encompass the presence of luminal irregularity, a segmental lumen 1.5 times larger than an adjacent one, or a visibly irregular luminal contour with a Z score exceeding 2.5 standard deviations^5–11^. In our previous investigations, which involved a sequential assessment of coronary artery lesions in 341 Taiwanese children with Kawasaki disease^12^, we found that 35% of KD patients experienced dilatation during the acute phase, 17.2% one month after the disease onset, and 10.2% during the two-month follow-up. Additionally, 4% showed persistent coronary artery lesions lasting for over one year.

Studies suggest that administering intravenous IVIG at a dose of 2 g/kg/day is highly successful in minimizing the likelihood of CAA in individuals with Kawasaki disease^13–16^. The approved therapy for the acute phase of Kawasaki disease, as recommended by the American Heart Association (AHA) and the American Academy of Pediatrics (AAP), includes the administration of IVIG with a single 2 g/kg dose given over 10-12 hours, along with oral high-dose aspirin at 80-100 mg/kg/day^17^. Although aspirin demonstrates notable anti-inflammatory effects in high doses and anti-platelet effects in low doses, there is no prospective study validating its role in diminishing the formation of CAL. According to two meta-analyses, high-dose aspirin alone does not effectively reduce the risk of CAA in KD; the reduction is predominantly reliant on the dose of IVIG^18, 19^. Several studies propose a strong correlation between the incidence of CAL and the dose of IVIG, indicating that the dosage of aspirin may not have a significant influence. This suggests that anti-platelet aspirin doses alone might be sufficient during the acute phase of KD. In a study by Hsieh et al., it was discovered that administering high-dose aspirin during the acute stage of KD did not affect the response rate to IVIG therapy, the duration of fever, or the occurrence of CAL when children were treated with a high IVIG dose (2 g/kg) in a single infusion^13^. In our retrospective analysis, which included 851 KD patients from two medical centers in Taiwan, we found no noteworthy distinctions between groups (with or without high-dose aspirin) with regard to gender, IVIG resistance, CAL formation, and the duration of hospitalization^16^. These retrospective findings imply that aspirin may offer minimal to no extra advantages when combined with IVIG therapy during the acute phase of KD.

At present, there is a shortage of well-conducted randomized controlled trials (RCTs) that can determine whether high-dose aspirin should continue to be a fundamental element of the treatment protocol for children diagnosed with KD. Furusho et al. conducted a study comparing the efficacy of IVIG and aspirin treatment against aspirin used alone in conjunction with IVIG^20^. According to Sanati et al., the use of high-dose aspirin does not have a substantial impact on preventing CAA in KD. In a prospective open-label trial conducted at a single site, administering the standard 2 g/kg/day IVIG without high-dose aspirin during acute-stage therapy did not elevate the risk of CAL in 62 patients with both typical and atypical KD^21^. Terai et al. discovered that the frequency of coronary abnormalities in KD was not affected by the aspirin dosage^19^. Furthermore, Chiang et al. performed a meta-analysis encompassing nine cohorts and a total of 12,182 children. The findings indicate that prescribing low-dose or omitting aspirin during the acute stage of KD might be linked to a lower occurrence of CAL^22^.

The risk associated with aspirin therapy in children with KD is generally considered low and appears to be comparable to risks reported in other situations. However, documented cases exist of severe side effects associated with aspirin in children undergoing KD treatment^16, 23–25^. Given the potential risks of drug toxicity and the limited robust evidence supporting the prevention of CAL formation, there is a necessity to reevaluate the role of high-dose aspirin during the acute stage of KD. To address this concern, we conducted a multi-center, prospective, parallel-group, open-label, non-inferiority randomized controlled trial to investigate whether high-dose aspirin, administered during the acute stage of KD, has beneficial effects in preventing the formation of CAL.

A recent investigative study, exploring the heterogeneity of Kawasaki disease, identified four unique patient clusters. These clusters were delineated according to subjective clinical features and laboratory results, resulting in the formation of the liver subgroup, band subgroup, node subgroup, and young subgroup^26^. It is noteworthy that these subgroups exhibited differences in their response to treatment and the outcomes of the disease, including the risk of coronary artery aneurysm and the rate of intravenous immunoglobulin resistance. Therefore, in this trial, an approach considering of these subgroups, following the recent proposal by the UCSD Burns group (liver/band/node/young), was also implemented. The goal was to compare the treatment effectiveness for each subgroup of Kawasaki disease between the standard and test arms, respectively.

## Methods

### Overall Study design

This study has been structured as a multi-center, prospective, randomized controlled, evaluator-blinded, non-inferiority trial with two parallel groups. The aim is to evaluate the effectiveness of intravenous immunoglobulin (IVIG) alone in comparison to IVIG combined with high-dose aspirin (80-100 mg/kg/day) as the primary treatment for Kawasaki disease (KD) during the acute stage. The clinical trial was conducted at five medical centers in Taiwan. Individuals under the age of 18 meeting the criteria for KD set by the American Heart Association (AHA) were eligible for enrollment in this study. Individuals with a fever of more than 10 days, previous treatment of steroids, biologics or were excluded. All patients were administered a high dose of intravenous immunoglobulin (IVIG) at 2 g/kg over a 12-hour duration, either with high-dose aspirin (80-100mg/kg/day) in group 1 (standard group) or without aspirin in group 2 (test group). Following the resolution of fever, low-dose aspirin (3-5 mg/kg/day) was prescribed for all KD patients for a period of 6-8 weeks or until normal echocardiography and inflammation laboratory data, in accordance with the guidelines provided by the AHA. The primary endpoint was identified as the formation of CAL at 6-8 weeks after enrollment.

This study was approved by the Institutional Review Board (IRB) at each site of the participating institutions, including Linkou Chang Gung Memorial Hospital, Kaohsiung Chang Gung Memorial Hospital, Taichung Veterans General Hospital, Kaohsiung Veterans General Hospital, and Tungs’ Taichung Metro Harbor Hospital. Informed consent form (ICF) were signed by the patient or a legal guardian.

### NCT registration

The study was registered at ClinicalTrials.gov with Identifier: NCT02951234 and Release Date: November 3 2016.

### Statistics method

All baseline characteristics were utilized to assess the comparability of both arms of the trial. The primary outcome evaluation and safety assessment were conducted using the intention-to-treat (ITT) population. Continuous variables, including age, gender, hospitalization duration (days), fever duration (days), height, weight, and clinical manifestations, were presented using either the mean ± standard deviation or frequency tables. Data analysis involved the use of the Chi-square test, independent t -test, Generalized Estimating Equation Model, and repeated measure analysis of variance (ANOVA) and non-inferiority comparison. The predetermined two-sided significance level for all statistical tests and confidence intervals was set at 5%.

## Results

### Patient enrollment and experimental design for the clinical trial

In this investigation, 152 patients diagnosed with Kawasaki Disease (KD) were enrolled (refer to Figure 1 for the enrollment flowchart). Participants were recruited from five medical centers in Taiwan. After excluding 18 cases, which included atypical KD cases or cases without completion of the study protocol, a final cohort of 134 KD patients was selected for further analysis. The schematic trial design is presented in supplementary Figure 1. The overall comparison between the test and standard groups is illustrated in supplementary Figure 1A: 69 patients were allocated to the standard group, receiving the standard treatment of intravenous immunoglobulin (2 g/kg) along with aspirin (80-100 mg/kg/day) until their fever subsided for 48 hours. The remaining 65 patients were assigned to the test group, receiving intravenous immunoglobulin (2 g/kg) alone. Majority of the fever was resolved within 24 hrs between the two groups after IVIG treatment. Following the initial treatment, both groups received a daily aspirin dose (3-5 mg/kg) for six weeks. The primary endpoint of this trial is the development of coronary artery lesions (CAL). Patients underwent 2D echocardiography at baseline, weeks 1, 2, 4, and 6-8, and again at 6 months. At 6 weeks and 6 months, the internal diameter and Z-score of the left and right main coronary arteries were calculated. CAL was defined for each age group: over 3.0 mm diameter for children under 5 years, over 4.0 mm for those 5 years or older, or any irregular luminal contour or Z-score exceeding 2.5. Throughout the study period, we also compared the treatment effect and prevention effect, as shown in supplementary Figure 1B and 1C, respectively. For the treatment effect, we followed CAL patients (Z score >2.5) at the acute phase to the endpoint. For the prevention effect, we followed non-CAL patients (Z score ≤2.5) at the acute phase to the endpoint.

**Figure 1:**
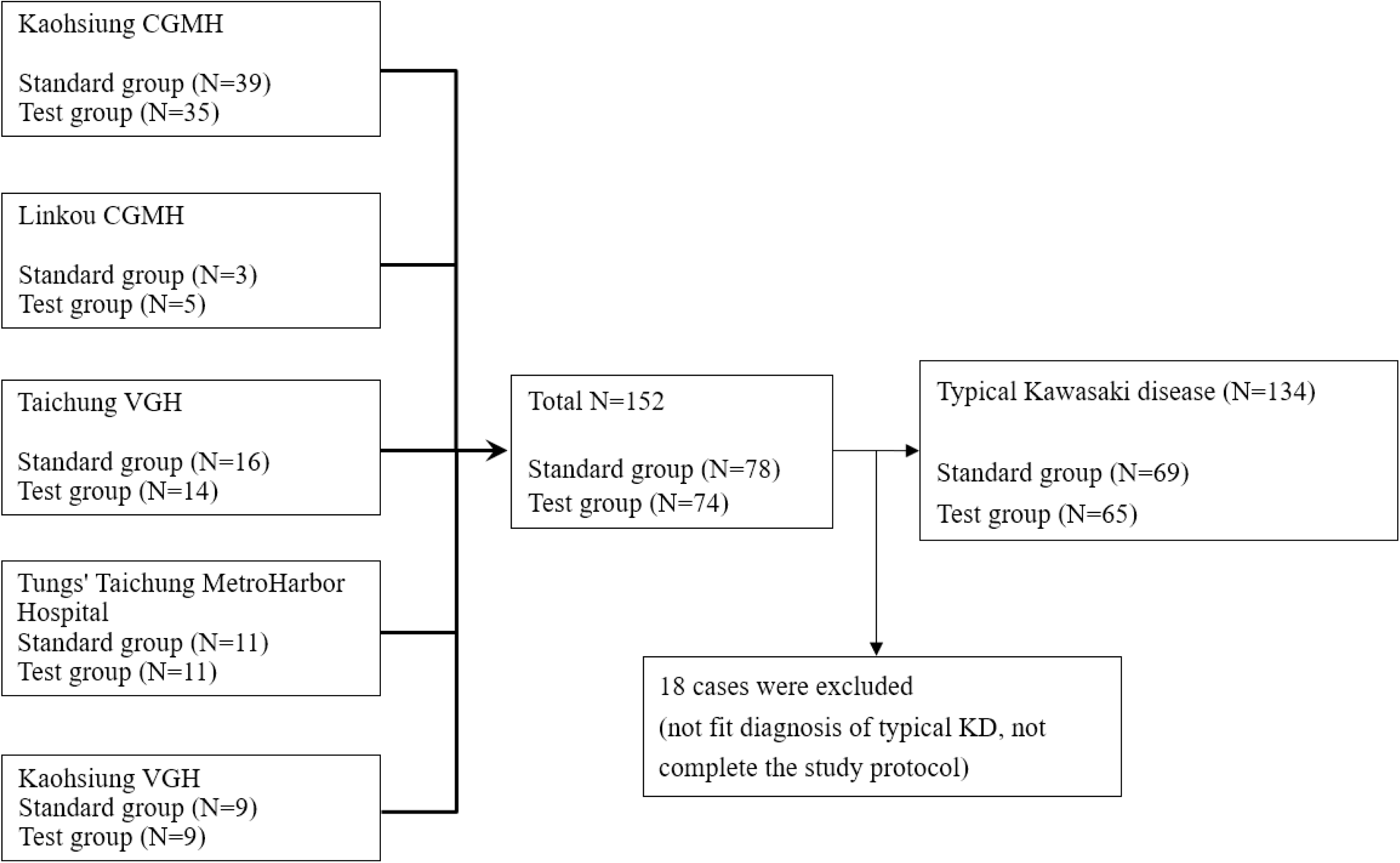
Consort diagram for patient enrollment. 152 KD patients were enrolled from five clinical centers. After removing 18 incomplete KD cases a final 134 KD patients were assigned into test and standard group for trial.

### Baseline Characteristics and Group Distribution of Participants in the Study

The percentage of males in the standard group (43%, n=30) and the test group (39%, n=25) did not exhibit a significant difference (P=0.933). Similarly, no statistically significant distinctions were observed between the groups in terms of mean weight (P=0.986) or height (P=0.969). Likewise, the mean ages of the two groups were comparable (P=0.464). Further analysis revealed no significant differences in baseline characteristics between the standard and test groups. Specifically, admission days (P=0.224), white blood cell counts (P=0.053), hemoglobin levels (P=0.138), platelet counts (P=0.074), GOT levels (P=0.668), and GPT levels (P=0.842) were all similar between the groups. The standard group exhibited significantly higher C-reactive protein levels at baseline compared to the test group (68.09 ± 76.66 vs. 41.86 ± 48.06 mg/L, P=0.021). Baseline coronary artery diameters and Z-scores in both the left main coronary artery (LMCA), right coronary artery (RCA), and left anterior descending artery (LAD) did not show significant differences between the groups (P=0.380, P=0.281, and P=0.788 for diameter; P=0.380, P=0.279, and P=0.674 for Z-score) (Table 1).

**Table 1.** Baseline Characteristics and Group Distribution of Participants in the Study.

| Variable | Mean ± SD | | | *P* |
| --- | --- | --- | --- | --- |
|  | Total<br>(n=134) | Standard Group<br>(n=69) | Test Group<br>(n=65) |  |
| Gender, n (%) |  |  |  |  |
| Male | 82 (61.2) | 43 (62.3) | 39 (60.0) | 0.783 |
| Female | 52 (38.8) | 26 (37.7) | 26 (40.0) |  |
| Age (years) | 1.77±1.31 | 1.85±1.43 | 1.69±1.17 | 0.464 |
| Weight (Kg) | 11.14±3.56 | 11.13±3.56 | 11.14±3.58 | 0.986 |
| Height (cm) | 81.83±13.13 | 81.87±13.85 | 81.79±12.44 | 0.969 |
| Admission (days) | 5.12±1.88 | 4.93±1.86 | 5.32±1.89 | 0.224 |
| White blood cell (WBC,<br>1000/μL) | 14.19±5.63 | 15.11±6.21 | 13.23±4.80 | 0.053 |
| Hemoglobin (g/dL) | 11.26±1.01 | 11.13±0.94 | 11.39±1.07 | 0.138 |
| Platelet (1000/μL) | 335.20±102.30 | 350.70±111.20 | 319.00±90.09 | 0.074 |
| CRP (mg/L) | 55.47±65.60 | 68.09±76.66 | 41.86±48.06 | 0.021* |
| GOT (U/L) | 95.38±143.70 | 90.20±120.80 | 101.30±166.80 | 0.668 |
| GPT (U/L) | 102.50±127.70 | 140.70±139.60 | 100.30±114.80 | 0.842 |
| LCMA diameter (mm) | 2.20±0.44 | 2.23±0.48 | 2.17±0.39 | 0.380 |
| LCMA Z score | 1.19±0.98 | 1.26±1.00 | 1.11±0.96 | 0.380 |
| LAD diameter (mm) | 1.75±0.42 | 1.74±0.44 | 1.76±0.40 | 0.788 |
| LAD Z score | 0.81±0.74 | 0.78±0.77 | 0.84±0.72 | 0.674 |
| RAC diameter (mm) | 1.91±0.43 | 1.87±0.44 | 1.95±0.41 | 0.281 |
| RAC Z score | 1.16±0.84 | 1.08±0.90 | 1.24±0.78 | 0.279 |

\* *P*<0.05
SD: Standard deviation, CRP: C-reactive protein, GOT: Glutamic oxaloacetic transaminase, GPT: Glutamic pyruvic transaminase, LMCA: Left main coronary artery, RCA: Right coronary artery, LAD: Left anterior descending

**Table 2.**
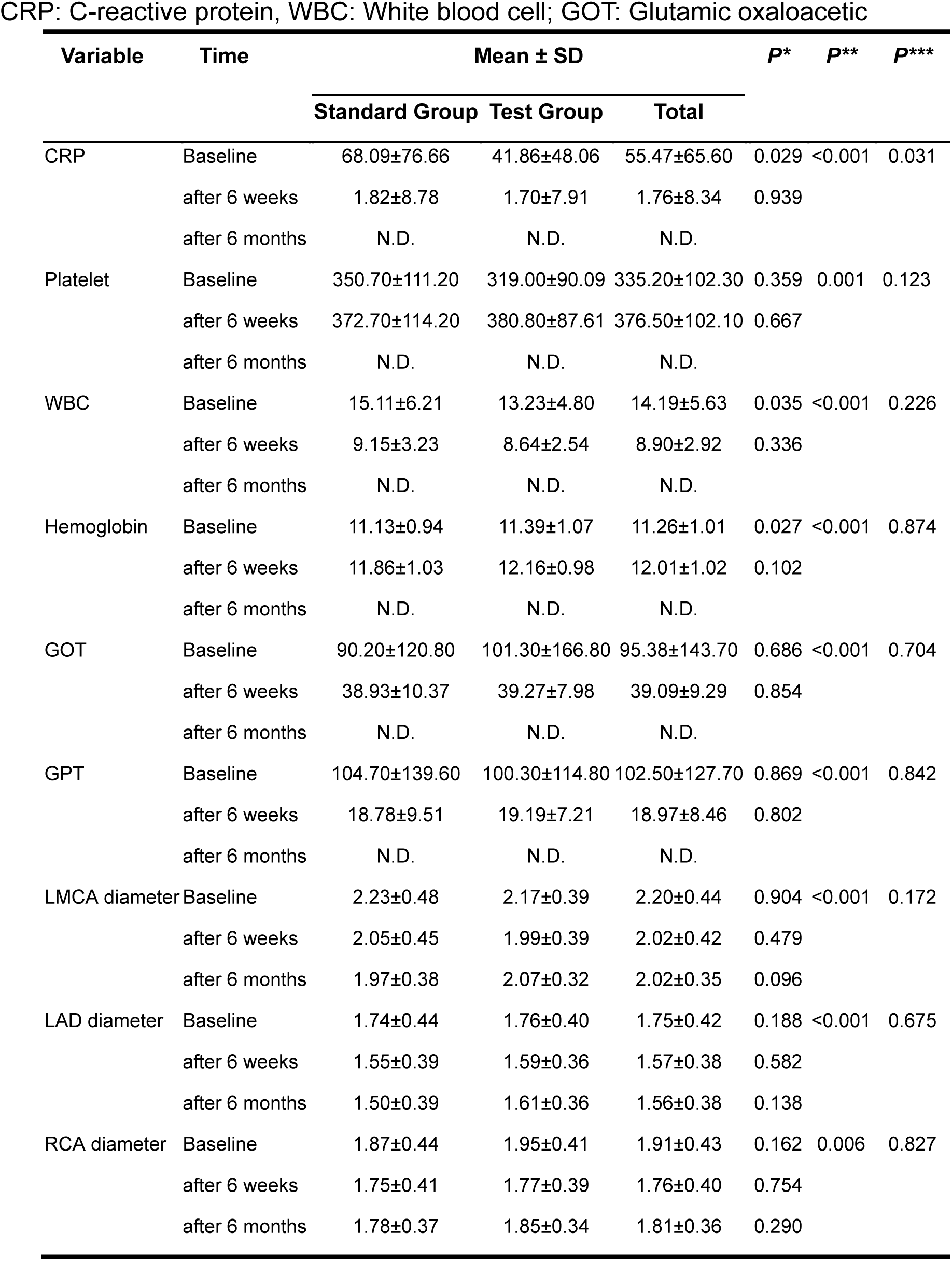
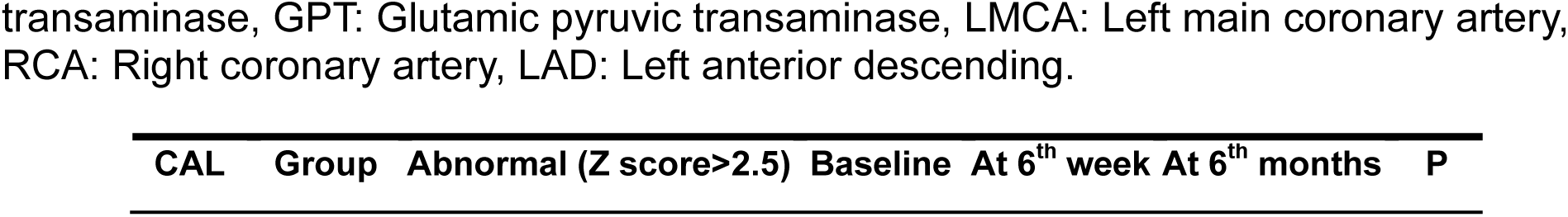
Analysis of variance of the effects of treatment in terms of the group, time, and their interaction (*Difference between two group, **Difference between the three times, ***Interaction effect between time and group)

**Table 3.**
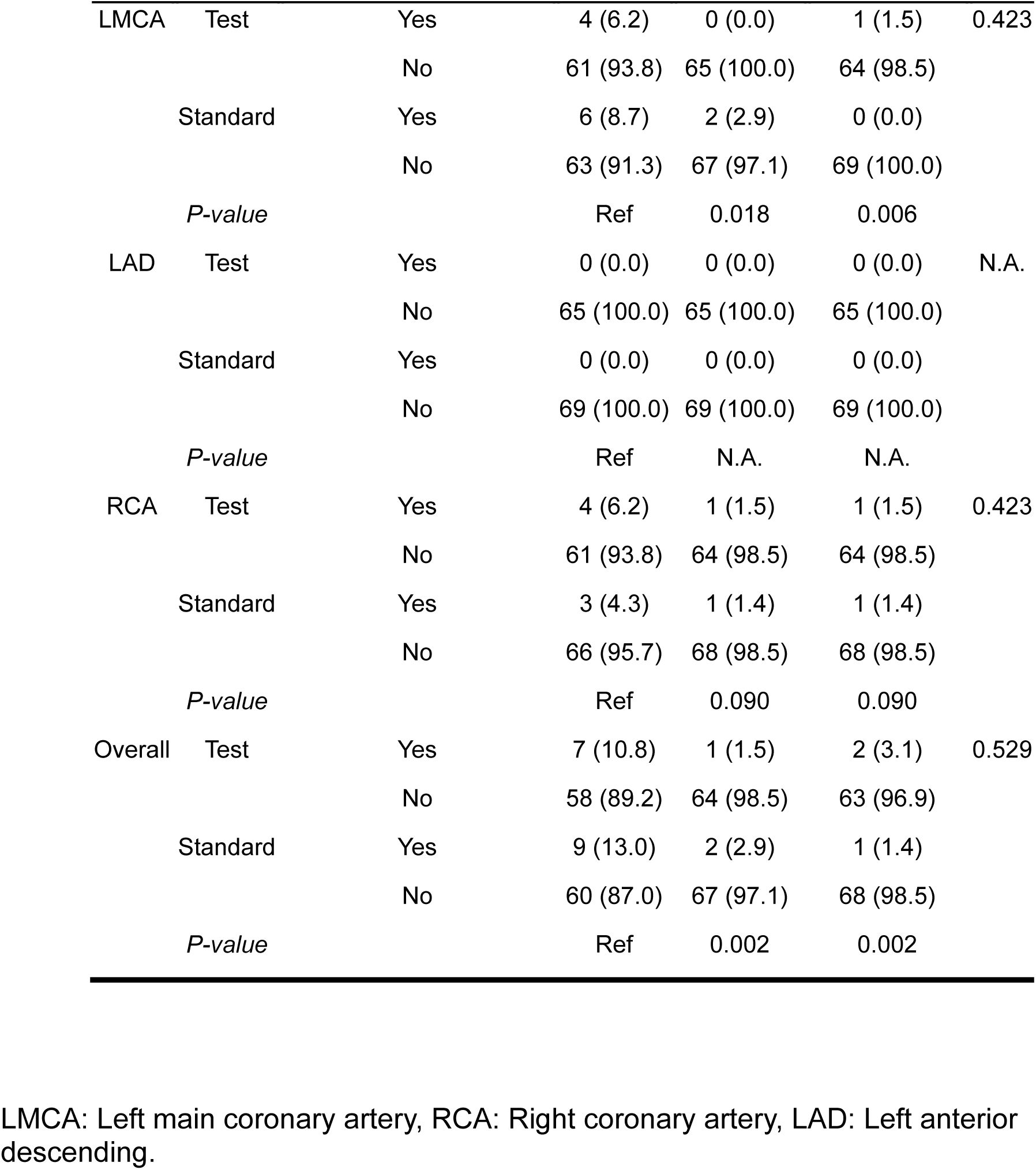
Comparison frequency of abnormal coronary arteries diameter in term of groups type and time-combined effect.

### Clinical Parameters Changes and Dynamics During the Study Period

Throughout the study, there was a noticeable decrease in C-reactive protein (CRP) levels. Starting from an average baseline of 55.47 mg/L (±65.60 SD), they decreased to just 1.76 mg/L (±8.34 SD) after 6 weeks. Although statistical analysis initially revealed a significant difference between the standard and test groups at baseline (P=0.029), this distinction disappeared by the 6-week mark (P=0.939). Overall, CRP levels exhibited a significant decline across the study duration (P<0.001), and interestingly, the specific pattern of this decline showed slight differences between the two groups (P=0.031 for interaction effect). Both groups demonstrated a general increase in the average platelet count after IVIG treatment compared to baseline levels. While the average platelet count changed significantly over time (P=0.001), there were no noticeable differences between the two groups at either baseline (P=0.359) or 6 weeks after treatment (P=0.667), and no interaction effect was observed between time and group type (P=0.123).

While white blood cell (WBC) levels exhibited a significant difference between the standard and test groups at baseline (P=0.035), this distinction disappeared by the 6th week (P=0.336). Overall, WBC levels displayed a significant decrease across the study duration (P<0.001), but the rate of decrease did not show a significant difference between the two groups (P=0.226). Hemoglobin levels increased significantly after 6 weeks in both groups (P<0.001). Although baseline levels differed significantly between the groups (P=0.020), this difference vanished by the 6th week (P=0.102). The rate of increase did not exhibit a significant difference between the groups (P=0.874). Two liver enzymes, glutamic oxaloacetic transaminase (GOT) and glutamic pyruvic transaminase (GPT), were monitored over time. Both enzymes showed a significant decrease throughout the study (P<0.001). Interestingly, their levels at baseline and after 6 weeks did not display a significant difference between the two groups (P=0.686 for GOT and P=0.809 for GPT), and the rate of decrease over time was also similar (P=0.704 for GOT and P=0.842 for GPT).

### Coronary Artery Changes and Frequency during study period

The diameter of the left main coronary artery (LMCA) significantly decreased across the study in both groups (p < 0.001), starting at 2.20 mm and reducing to 2.02 mm after 6 months. The rate of reduction was similar between the groups at each time point (P=0.904 for baseline, P=0.479 for 6th week, and P=0.096 for 6th month), and there was no significant interaction effect between time and groups (P=0.172). Similarly, the left anterior descending artery (LAD) exhibited a significant narrowing in both groups (p < 0.001), starting at 1.75 mm and decreasing to 1.56 mm after 6 months. Once again, the pattern of reduction was comparable between the groups (P=0.188 for baseline, P=0.582 for 6th week, and P=0.138 for 6th month). However, no interaction effect was observed between time and groups (P=0.675). Likewise, the right coronary artery (RCA) narrowed significantly over time (P = 0.006), from 1.91 mm to 1.81 mm after 6 months, with no noticeable differences in the rate of reduction between the groups (P=0.162 for baseline, P=0.754 for 6th week, and P=0.290 for 6th month), and there was no significant interaction between time and RCA in the groups (P=0.827).

Overall, at baseline, 10.8% of patients in the test group and 13.0% in the standard group presented abnormal coronary artery diameter (Z score >2.5) in at least one of the three coronary arteries. By the 6th month, these percentages reduced to 3.1% and 1.4%, respectively. No significant differences were observed between the groups regarding the frequency of coronary artery abnormalities during the study period (P = 0.529). The frequency of coronary artery lesions exhibited a significant decrease compared to the baseline, starting from the 6th week (P=0.002) through the end of the study period (6th month P=0.002). Similarly, no significant differences were observed between the two groups on either LMCA or RCA (detailed analysis in the supplementary appendix).

As for the non-inferiority comparison between the two groups, we consider a difference in CAL rate of 10% or less as clinically insignificant and the noninferiority margin was set at 10%. The CAL rate was 12.3% (8/65; 95% confidence interval: 5.5% to 22.8%) in the test group and 15.9% (11/69; 95% confidence interval: 8.2% to 26.7%) in the standard care group. The one-sided 97.5% confidence interval for the CAL difference between the test and standard care group was [-100%, 8.1%]. The upper end of the confidence interval was below the noninferiority margin of 10%. Therefore, we can conclude with 97.5% confidence that the CAL rate in the test group was no higher than that in the standard care group by 10% and the noninferiority was established in current cohort size of patient numbers in both standard and test groups.

In addition to assessing CAL as the primary outcome, we also investigated IVIG resistance as a secondary outcome. Table S1 indicates that both groups had 3 patients each with IVIG resistance. There were no significant differences in the rates of IVIG resistance between the groups (P=0.940).

### Treatment and Prevention Effect on Coronary Artery Changes and Frequency during Study Period between Two Groups

Throughout the study duration, we noted that most CAL cases were detected during the acute phase. However, some patients were initially normal during the acute phase but later developed CAL at 6 weeks or 6 months, even after receiving IVIG or IVIG plus high-dose treatment. To compare the treatment effect between the test and standard groups, we conducted statistical analysis of CAL frequency exclusively on the CAL patients identified during the acute phase.

As depicted in Table 4, overall, at the baseline, 10.8% of patients in the test group and 13.0% in the standard group presented abnormal coronary artery diameter (Z score >2.5) in at least one of the three coronary arteries. By the 6th month, all patients had recovered from CAL. No significant differences were observed between the groups concerning the frequency of abnormal coronary artery abnormalities during the study period (P = 0.423). The frequency of coronary artery lesions exhibited a significant decrease compared to the baseline, starting from the 6th week (P=0.001) through the end of the study period (6th month P<0.001). Similarly, no significant treatment differences were observed between the two groups on either LMCA or RCA (detailed analysis in the supplementary appendix).

**Table 4.**
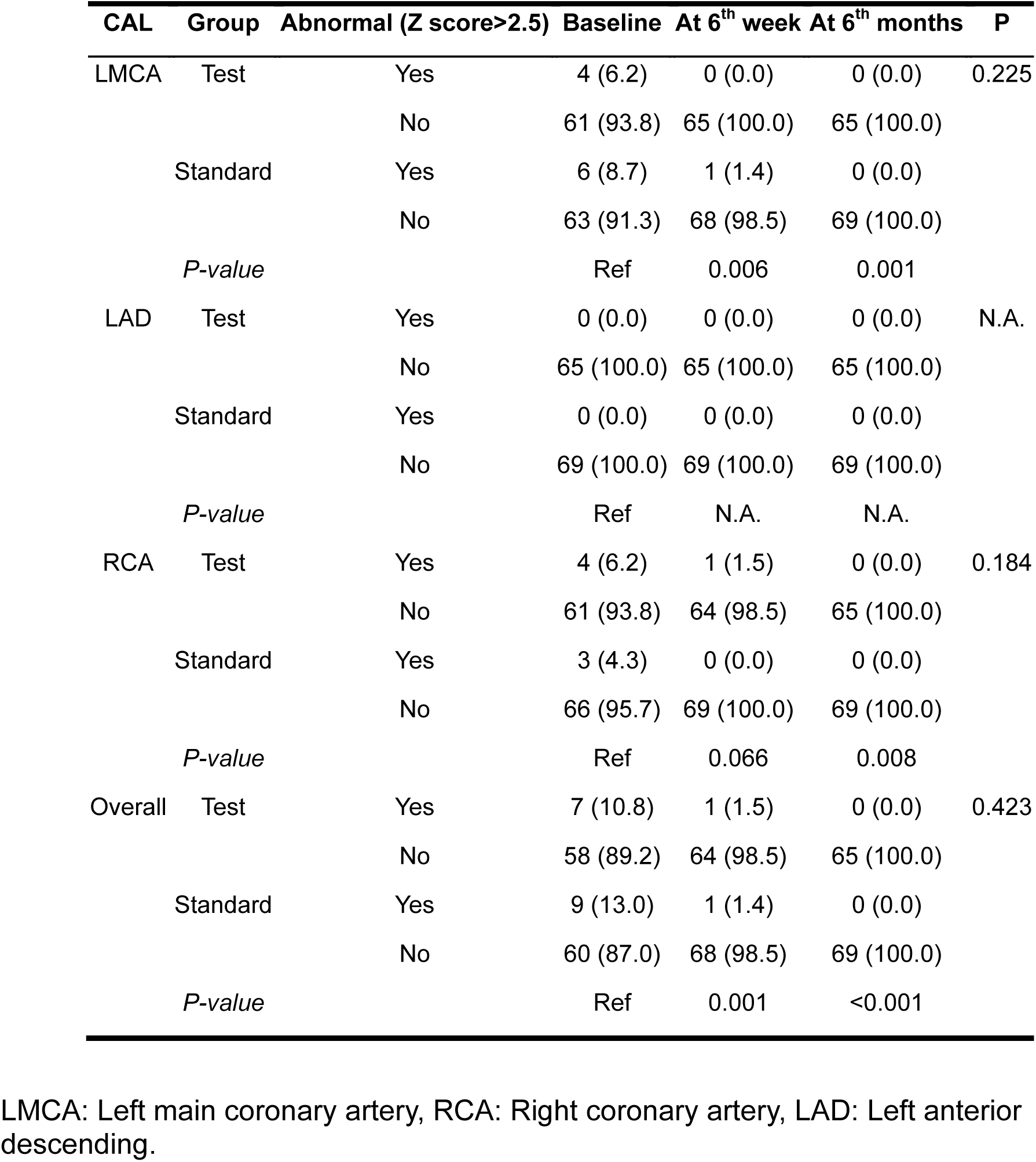
Comparison frequency of abnormal coronary arteries diameter in term of groups type and time-treatment effect.

| CAL | Group | Abnormal (Z score>2.5) | Baseline | At 6<sup>th</sup> week | At 6<sup>th</sup> months | P |
| --- | --- | --- | --- | --- | --- | --- |
| LMCA | Test | Yes | 4 (6.2) | 0 (0.0) | 0 (0.0) | 0.225 |
|  |  | No | 61 (93.8) | 65 (100.0) | 65 (100.0) |  |
|  | Standard | Yes | 6 (8.7) | 1 (1.4) | 0 (0.0) |  |
|  |  | No | 63 (91.3) | 68 (98.5) | 69 (100.0) |  |
|  | *P-value* |  | Ref | 0.006 | 0.001 |  |
| LAD | Test | Yes | 0 (0.0) | 0 (0.0) | 0 (0.0) | N.A. |
|  |  | No | 65 (100.0) | 65 (100.0) | 65 (100.0) |  |
|  | Standard | Yes | 0 (0.0) | 0 (0.0) | 0 (0.0) |  |
|  |  | No | 69 (100.0) | 69 (100.0) | 69 (100.0) |  |
|  | *P-value* |  | Ref | N.A. | N.A. |  |
| RCA | Test | Yes | 4 (6.2) | 1 (1.5) | 0 (0.0) | 0.184 |
|  |  | No | 61 (93.8) | 64 (98.5) | 65 (100.0) |  |
|  | Standard | Yes | 3 (4.3) | 0 (0.0) | 0 (0.0) |  |
|  |  | No | 66 (95.7) | 69 (100.0) | 69 (100.0) |  |
|  | *P-value* |  | Ref | 0.066 | 0.008 |  |
| Overall | Test | Yes | 7 (10.8) | 1 (1.5) | 0 (0.0) | 0.423 |
|  |  | No | 58 (89.2) | 64 (98.5) | 65 (100.0) |  |
|  | Standard | Yes | 9 (13.0) | 1 (1.4) | 0 (0.0) |  |
|  |  | No | 60 (87.0) | 68 (98.5) | 69 (100.0) |  |
|  | *P-value* |  | Ref | 0.001 | <0.001 |  |

LMCA: Left main coronary artery, RCA: Right coronary artery, LAD: Left anterior descending.

Following treatment effect, we investigated the preventive impact on the frequency of Coronary Artery Lesions (CAL) over the study duration for patients who developed CAL later after IVIG or IVIG plus high-dose aspirin treatment. We performed the same statistical analysis for the comparison of CAL frequencies, as illustrated in Supplementary Table 2. No statistical difference was identified between the standard and test groups. However, a minimal number of newly developed CAL cases were observed after treatment: 2 in the test group and 1 in the standard group.

## Discussion

Preventing coronary artery lesions (CALs) is crucial as it represents the most severe complication of Kawasaki disease (KD). Therefore, determining the ideal aspirin dosage to prevent this complication is of paramount importance. Aspirin, a salicylate drug, works by inhibiting the production of prostaglandins, which are involved in the inflammatory response. By exerting anti-inflammatory and antiplatelet effects, aspirin is thought to play a role in suppressing the inflammatory response and reducing the risk of blood clots in Kawasaki disease.

Our previous study demonstrated that the use of low-dose aspirin in the initial treatment of children with KD was not linked to fever recurrence or the formation of CAL. Additionally administering high-dose aspirin during the acute stage of KD did not yield any benefits in terms of inflammation or improved treatment outcomes^16^. This study represents the first multi-center randomized controlled trial investigating the efficacy of high-dose aspirin or intravenous immunoglobulin (IVIG) during the acute stage of KD. In this clinical trial, we assessed the impact of discontinuing high-dose aspirin (80–100 mg/kg/day) on the occurrence of CAL during the acute phase treatment of KD in both the test and standard groups. The findings revealed that the elimination of high-dose aspirin did not yield a significant effect on CAL incidence. No significant differences were observed between the groups in terms of the frequency of abnormal coronary artery abnormalities during the study period (P = 0.529). Through a statistically robust non-inferiority comparison, we concluded with 97.5% confidence that the CAL rate in the test group was not higher than that in the standard care group by 10%, establishing noninferiority with clinical insignificance.

Evaluating the overall effectiveness of IVIG treatment entails considering both its treatment effect (the number of cases successfully treated) and prevention effect (the prevention of new cases). Our analysis revealed no statistically significant differences in either the treatment effect (P=0.423) or prevention effect (P>0.999) between these two groups, despite the occurrence of very limited newly developed CAL cases. Conclusions regarding the prevention effect may require a larger cohort to ensure robust findings.

Furthermore, we performed an analysis to compare the effectiveness in four subgroups of KD, which were categorized based on subjective clinical features and laboratory results. Within each of these individual clusters or subgroups, encompassing two clusters with distinct characteristics, no significant difference was observed in the reduction of CAL rates at the 6^th^ week or 6^th^ month after treatment during the acute phase between the test and standard groups.

The primary purpose of administering high-dose aspirin was to elicit anti-inflammatory effects. As a result, we conducted an analysis comparing efficacy in subgroups classified based on the acute phase levels of two inflammatory biomarkers: C-reactive protein (CRP) and platelet count. Consistent with findings in other subgroups, we identified no significant distinction between the test and standard groups in either the low (normal range) subgroups for CRP or platelet count, nor in the high (abnormal range) subgroups. The analysis of these subgroups indicates that the administration of IVIG treatment alone, without the addition of high-dose aspirin, does not increase the risk of CAL six weeks or later after the initial treatment. Furthermore, this approach has wider relevance for KD patients with varied demographic and clinical characteristics.

In conclusion, in our trial, high-dose aspirin does not play a significant role in managing coronary artery lesions in KD patients. Administering the standard 2 g/kg/day IVIG without high-dose aspirin (80–100 mg/kg/day) during the acute phase therapy for KD does not increase the risk of coronary artery lesions, which are a primary cause of morbidity and mortality in KD patients. Results from comparison analysis indicate the non-inferiority between two groups. Therefore, addition of high-dose aspirin during initial IVIG treatment is not statistically significant or clinically meaningful.

## Data Availability

The data that support the findings of this study are available from the corresponding author on reasonable request. Participant data without names and identifiers will be made available after approval from the corresponding author and Chang Gung Memorial Hospital. After publication of study findings, the data will be available for others to request.

## Contributors

H-CK, HJC and XBL contributed to the study conception and design. M-CL, C-CK, K-PW, YD, ZH, and LT contributed to the statistical analyses, data entry, and verification. All authors were involved in the study implementation, data acquisition, and interpretation of the results. All authors critically reviewed the manuscript, approved the final version for submission, and agree to be accountable for all aspects of this work.

## Declaration of interests

All the authors hereby declare to have no financial interests to disclose regarding this article.

## Acknowledgements

The authors thank the patients and their carers who participated in this study. This study received funding from the following grants: CMRPG8J1151 CPRPG8F0791 CPRPG8H0051-2 CMRPG8M1431 CMRPG8M1421 and CMRPG8L1241-2 from Chang Gung Memorial Hospital and the NSTC 112-2314-B-182-032-MY3 from National Science and Technology Council of Taiwan to Dr. Ho; and 1R41TR004351-01 from US National Institute of Health to Dr. Ling. Although these institutes provided financial support, they had no influence on the way in which we collected analyzed or interpreted the data or wrote this manuscript.

## Notes

### Competing Interest Statement

The authors have declared no competing interest.

### Clinical Trial

NCT02951234

## Reference

1. Burns JC, Glode MP. Kawasaki syndrome. Lancet. 2004;364(9433):533–44. doi: 10.1016/S0140-6736(04)16814-1. PubMed PMID: 15302199.

2. Eleftheriou D, Levin M, Shingadia D, Tulloh R, Klein NJ, Brogan PA. Management of Kawasaki disease. Arch Dis Child. 2014;99(1):74–83. Epub 20131025. doi: 10.1136/archdischild-2012-302841. PubMed PMID: 24162006; PMCID: PMC3888612.

3. Liang CD, Kuo HC, Yang KD, Wang CL, Ko SF. Coronary artery fistula associated with Kawasaki disease. Am Heart J. 2009;157(3):584–8. Epub 20090203. doi: 10.1016/j.ahj.2008.11.020. PubMed PMID: 19249434.

4. Newburger JW, Takahashi M, Gerber MA, Gewitz MH, Tani LY, Burns JC, Shulman ST, Bolger AF, Ferrieri P, Baltimore RS, Wilson WR, Baddour LM, Levison ME, Pallasch TJ, Falace DA, Taubert KA, Committee on Rheumatic Fever E, Kawasaki D, Council on Cardiovascular Disease in the Y, American Heart A, American Academy of P. Diagnosis, treatment, and long-term management of Kawasaki disease: a statement for health professionals from the Committee on Rheumatic Fever, Endocarditis and Kawasaki Disease, Council on Cardiovascular Disease in the Young, American Heart Association. Circulation. 2004;110(17):2747–71. doi: 10.1161/01.CIR.0000145143.19711.78. PubMed PMID: 15505111.

5. Akagi T, Rose V, Benson LN, Newman A, Freedom RM. Outcome of coronary artery aneurysms after Kawasaki disease. J Pediatr. 1992;121(5 Pt 1):689–94. doi: 10.1016/s0022-3476(05)81894-3. PubMed PMID: 1432415.

6. Shulman ST, De Inocencio J, Hirsch R. Kawasaki disease. Pediatr Clin North Am. 1995;42(5):1205–22. doi: 10.1016/s0031-3955(16)40059-3. PubMed PMID: 7567192.

7. Yu HR, Kuo HC, Sheen JM, Wang L, Lin IC, Wang CL, Yang KD. A unique plasma proteomic profiling with imbalanced fibrinogen cascade in patients with Kawasaki disease. Pediatr Allergy Immunol. 2009;20(7):699–707. Epub 20090112. doi: 10.1111/j.1399-3038.2008.00844.x. PubMed PMID: 19170925.

8. Wu MT, Hsieh KS, Lin CC, Yang CF, Pan HB. Images in cardiovascular medicine. Evaluation of coronary artery aneurysms in Kawasaki disease by multislice computed tomographic coronary angiography. Circulation. 2004;110(14):e339. doi: 10.1161/01.CIR.0000143374.80173.EF. PubMed PMID: 15466653.

9. Weng KP, Ho TY, Chiao YH, Cheng JT, Hsieh KS, Huang SH, Ou SF, Liu KH, Hsu CJ, Lu PJ, Hsiao M, Ger LP. Cytokine genetic polymorphisms and susceptibility to Kawasaki disease in Taiwanese children. Circ J. 2010;74(12):2726–33. Epub 20101030. doi: 10.1253/circj.cj-10-0542. PubMed PMID: 21048327.

10. Weng KP, Hsieh KS, Ho TY, Huang SH, Lai CR, Chiu YT, Huang SC, Lin CC, Hwang YT, Ger LP. IL-1B polymorphism in association with initial intravenous immunoglobulin treatment failure in Taiwanese children with Kawasaki disease. Circ J. 2010;74(3):544–51. Epub 20100118. doi: 10.1253/circj.cj-09-0664. PubMed PMID: 20081319.

11. Weng KP, Hsieh KS, Hwang YT, Huang SH, Lai TJ, Yuh YS, Hou YY, Lin CC, Huang SC, Chang CK, Lin MW, Ger LP. IL-10 polymorphisms are associated with coronary artery lesions in acute stage of Kawasaki disease. Circ J. 2010;74(5):983–9. Epub 20100326. doi: 10.1253/circj.cj-09-0801. PubMed PMID: 20339193.

12. Kuo HC, Yu HR, Juo SH, Yang KD, Wang YS, Liang CD, Chen WC, Chang WP, Huang CF, Lee CP, Lin LY, Liu YC, Guo YC, Chiu CC, Chang WC. CASP3 gene single-nucleotide polymorphism (rs72689236) and Kawasaki disease in Taiwanese children. J Hum Genet. 2011;56(2):161–5. Epub 20101216. doi: 10.1038/jhg.2010.154. PubMed PMID: 21160486.

13. Hsieh KS, Weng KP, Lin CC, Huang TC, Lee CL, Huang SM. Treatment of acute Kawasaki disease: aspirin’s role in the febrile stage revisited. Pediatrics. 2004;114(6):e689–93. Epub 20041115. doi: 10.1542/peds.2004-1037. PubMed PMID: 15545617.

14. Lee G, Lee SE, Hong YM, Sohn S. Is high-dose aspirin necessary in the acute phase of kawasaki disease? Korean Circ J. 2013;43(3):182–6. Epub 20130331. doi: 10.4070/kcj.2013.43.3.182. PubMed PMID: 23613695; PMCID: PMC3629244.

15. Saulsbury FT. Comparison of high-dose and low-dose aspirin plus intravenous immunoglobulin in the treatment of Kawasaki syndrome. Clin Pediatr (Phila). 2002;41(8):597–601. doi: 10.1177/000992280204100807. PubMed PMID: 12403377.

16. Kuo HC, Lo MH, Hsieh KS, Guo MM, Huang YH. High-Dose Aspirin is Associated with Anemia and Does Not Confer Benefit to Disease Outcomes in Kawasaki Disease. PLoS One. 2015;10(12):e0144603. Epub 20151210. doi: 10.1371/journal.pone.0144603. PubMed PMID: 26658843; PMCID: PMC4686074.

17. McCrindle BW, Rowley AH, Newburger JW, Burns JC, Bolger AF, Gewitz M, Baker AL, Jackson MA, Takahashi M, Shah PB, Kobayashi T, Wu MH, Saji TT, Pahl E, American Heart Association Rheumatic Fever E, Kawasaki Disease Committee of the Council on Cardiovascular Disease in the Y, Council on C, Stroke N, Council on Cardiovascular S, Anesthesia, Council on E, Prevention. Diagnosis, Treatment, and Long-Term Management of Kawasaki Disease: A Scientific Statement for Health Professionals From the American Heart Association. Circulation. 2017;135(17):e927–e99. Epub 20170329. doi: 10.1161/CIR.0000000000000484. PubMed PMID: 28356445.

18. Durongpisitkul K, Gururaj VJ, Park JM, Martin CF. The prevention of coronary artery aneurysm in Kawasaki disease: a meta-analysis on the efficacy of aspirin and immunoglobulin treatment. Pediatrics. 1995;96(6):1057–61. PubMed PMID: 7491221.

19. Terai M, Shulman ST. Prevalence of coronary artery abnormalities in Kawasaki disease is highly dependent on gamma globulin dose but independent of salicylate dose. J Pediatr. 1997;131(6):888–93. doi: 10.1016/s0022-3476(97)70038-6. PubMed PMID: 9427895.

20. Furusho K, Kamiya T, Nakano H, Kiyosawa N, Shinomiya K, Hayashidera T, Tamura T, Hirose O, Manabe Y, Yokoyama T, et al. Intravenous gamma-globulin for Kawasaki disease. Acta Paediatr Jpn. 1991;33(6):799–804. doi: 10.1111/j.1442-200x.1991.tb02611.x. PubMed PMID: 1801560.

21. Sanati F, Bagheri M, Eslami S, Khalooei A. Evaluation of high-dose aspirin elimination in the treatment of Kawasaki disease in the incidence of coronary artery aneurysm. Ann Pediatr Cardiol. 2021;14(2):146–51. Epub 20210416. doi: 10.4103/apc.APC_206_20. PubMed PMID: 34103852; PMCID: PMC8174624.

22. Chiang MH, Liu HE, Wang JL. Low-dose or no aspirin administration in acute-phase Kawasaki disease: a meta-analysis and systematic review. Arch Dis Child. 2021;106(7):662–8. Epub 20201110. doi: 10.1136/archdischild-2019-318245. PubMed PMID: 33172886.

23. Matsubara T, Mason W, Kashani IA, Kligerman M, Burns JC. Gastrointestinal hemorrhage complicating aspirin therapy in acute Kawasaki disease. J Pediatr. 1996;128(5 Pt 1):701-3. doi: 10.1016/s0022-3476(96)80140-5. PubMed PMID: 8627447.

24. Wei CM, Chen HL, Lee PI, Chen CM, Ma CY, Hwu WL. Reye’s syndrome developing in an infant on treatment of Kawasaki syndrome. J Paediatr Child Health. 2005;41(5-6):303–4. doi: 10.1111/j.1440-1754.2005.00617.x. PubMed PMID: 15953335.

25. Lee JH, Hung HY, Huang FY. Kawasaki disease with Reye syndrome: report of one case. Zhonghua Min Guo Xiao Er Ke Yi Xue Hui Za Zhi. 1992;33(1):67–71. PubMed PMID: 1626454.

26. Wang H, Shimizu C, Bainto E, Hamilton S, Jackson HR, Estrada-Rivadeneyra D, Kaforou M, Levin M, Pancheri JM, Dummer KB, Tremoulet AH, Burns JC. Subgroups of children with Kawasaki disease: a data-driven cluster analysis. Lancet Child Adolesc Health. 2023;7(10):697–707. Epub 20230817. doi: 10.1016/S2352-4642(23)00166-9. PubMed PMID: 37598693; PMCID: PMC10756500.

